# A systematic review of the effect of renal replacement therapy on the pharmacokinetics of co-amoxiclav

**DOI:** 10.1101/2024.06.19.24309187

**Authors:** Sarraa Al-Mahdi, Jignna Patel, James Sweatman, Robert Oakley, Reya V. Shah, Joseph F. Standing, Dagan Lonsdale

## Abstract

1.

**Background & Aim:** Co-amoxiclav is a commonly used antibiotic. Although, the dose administered during renal replacement therapy may be subtherapeutic. This study aims to describe the current literature on the pharmacokinetics and pharmacodynamics of co-amoxiclav in patientss undergoing renal replacement therapy.

**Method:** We carried out a systematic review of the available literature in MEDLINE, Embase, Pubmed, and Google Scholar from inception to Oct 2023. Studies were included if they reported pharmacokinetic data on adults given amoxicillin or clavulanic acid during renal replacement therapy.

**Results:** Seven studies were identified which were published between 1984 to 2021. Variability was observed in the participant characteristics within the studies, the renal replacement therapy settings, the drug exposure, drug assay methods, and the analysis of the pharmacokinetic parameters.

**Conclusion:** Further pharmacokinetic-pharmacodynamic studies are needed on co-amoxiclav during renal replacement therapy.

## 2. Introduction

### Review Question

What are the pharmacokinetics of co-amoxiclav in patients undergoing renal replacement therapy?

### Introduction

Antibiotics are the cornerstone of therapy for the treatment of bacterial infections. Co-amoxiclav is a combination of two separate drugs: amoxicillin and clavulanic acid. Amoxicillin is a penicillin derivative that has activity against both gram-positive and gram-negative bacteria. Amoxicillin is a bactericidal agent that targets and kills bacteria by inhibiting the biosynthesis of the peptidoglycan layer of the bacterial cell wall [1].

Bacterial resistance to amoxicillin is frequently caused by the production of beta-lactamase which inactivates amoxicillin. Calvulanic acid functions as an inhibitor of beta-lactamase, it has no antimicrobial activity of its own. It works by preventing bacterial destruction of beta-lactams by bacteria consequently restoring the antimicrobial effects of amoxicillin [1].

Co-amoxiclav is used commonly to treat infections caused by beta-lactamase producing strains, including those affecting the respiratory tract, bone, joint, genito-urinary, and abdominal areas [2].

The molecular weight of amoxicillin is 365.4 daltons, clavulanic acid is 199.2 daltons. The percentage of protein binding is low 20% for amoxicillin and 25% for clavulanic acid respectively. The percentage of amoxicillin excreted unchanged in urine is 60% and for clavulanic acid 40% respectively. The volume of distribution (Vd) of amoxicillin in healthy adults is 0.3 L/kg, and the Vd of clavulanic acid is 0.3 L/kg. The half-life of amoxicillin in healthy adults is 1-1.5h and in end-stage renal failure, the half-life is between 7 to 20 h. The half-life of clavulanic acid is 1h and in end-stage renal failure, the half-life is between 3-4 h. [3]

The free time above the minimum inhibitory concentration (fT>MIC) is considered to be the major determinant of efficacy for amoxicillin. The MIC susceptibility breakpoint for amoxicillin as stated by the European Committee on Antimicrobial Susceptibility Testing (EUCAST) ranges between 0.25 to 8 mg/L. Clavulanic acid has a fixed breakpoint of 2mg/L [4].

We know that giving the right dose of antibiotics is really important and co-amoxiclav is excreted by the kidneys. When a patient’s kidneys fail, the dose of co-amoxiclav is often reduced to prevent adverse effects. Severe infections can cause organ damage to the kidneys necessitating renal replacement therapy in 10% of septic patients [5]. Currently, we do not know whether modern renal replacement machines clear co-amoxiclav. Consequently, lower doses of co-amoxiclav are given to these patients [3].

A recent multinational study of patients with sepsis who were receiving renal replacement therapy has highlighted that this approach is likely to be wrong. They showed that almost three-quarters of patients received doses of antibiotics that were too low [3, 6] Underdosing could result in treatment failure and increased antibiotic resistance while overdosing could result in drug toxicity [7].

In addition, patients who have chronic kidney disease and are dependent on regular renal replacement therapy are immunocompromised; they have a higher incidence of antimicrobial resistance and will need frequent treatment using antimicrobials [8].

RRT offers different modes of dialysis using diffusion, convection, and ultrafiltration with different effluent dosages. There are differences in the duration of RRT given as continuous renal replacement therapy, intermittent dialysis, or slow extended dialysis. The type of filter and pore size impact the clearance of drug molecules. The effluent rate and the sieving coefficient of the drug also impact the clearance of a drug [8]. These RRT factors could impact the optimal dose of co-amoxiclav.

The lack of consensus guidance for co-amoxiclav dosage has resulted in variability in dosing adjustments in clinical practice, with different and sometimes conflicting resources being referenced [1, 3, 9].

Pharmacokinetic characteristics of co-amoxiclav in RRT are not required for licensing, little is known about safe and efficacious co-amoxiclav dosing in these circumstances.

To provide more clarity on the available evidence to inform clinical practice, this systematic review focuses on establishing what has been published regarding the pharmacokinetics and pharmacodynamics of co-amoxiclav when given to patients undergoing renal replacement therapy. This information is important to better inform and subsequently improve co-amoxiclav treatment for patients needing renal replacement therapy.

We aim to identify the clearance, volume of distribution, and the recommended dosage for co-amoxiclav during RRT.

## 3 Methods

### 3.1 Data Source and Searches

This systematic review was guided by the Preferred Reported Items for Systematic Reviews and Meta-Analysis (PRISMA) guidelines [10]. The protocol for this systematic review was registered on PROSPERO (CDR42023469918). The search strategy is available in Appendix 1.

We searched the following databases from inception to 4 Oct 2023: MEDLINE, Embase, Pubmed, and Google Scholar. Two reviewers (SAM and JP) conducted the search independently. The search terms were: amoxicillin; clavulanic; dialysis; haemodialysis or haemofiltration or heamodiafiltration; continuous renal replacement or continuous renal replacement therapy or CRRT; continuous venovenous heamodiafiltration; renal replacement or RRT; intermittent haemodialysis or IHD, sustained low efficiency dialysis or SLED; drug monitoring; dose response relationship; therapeutic drug monitoring or TDM; pharmaco* or PK or PKPD; pharmacokinetics, pharmacodynamics. The search was limited to English language, humans, and adults aged >16. There were no restrictions on the year of publication. The titles and abstracts were independently screened; the full text of relevant papers were reviewed according to the selection criteria. If necessary, a third investigator (DL) was available to resolve ambiguities or disagreements regarding the selection of studies. The reference list of papers included and relevant review papers were manually searched for further relevant papers that meet the inclusion criteria.

### 3.2 Study Selection

We included case reports, case-controlled studies, cohort studies, randomised controlled trials, systematic reviews, meta-analysis, and conference abstracts. Qualitative research, literature reviews, and other texts were excluded. The study was eligible for inclusion if it met the following criteria: (1) includes amoxicillin; (2) includes clavulanic acid; (3) patients must be undergoing renal replacement therapy (RRT) via the following modes: continuous venovenous haemofiltration (CVVHF), continuous venovenous haemodialysis (CVVHD), continuous venovenous haemodiafiltration (CVVHDF), sustained low efficiency dialysis (SLED), haemodialysis (HD) or high flux haemodialysis; (4) peer-reviewed; (5) available in the English language. Studies were excluded if they met the following criteria: (1) Computational or model based studies not based on patient data (2) Non-human studies (3) Subjects aged <16 (4) Insufficient pharmacokinetic data on study drug’s clearance, Vd or half-life. Fig 1 shows the PRISMA flow chart for study inclusion.

**Fig 1:**
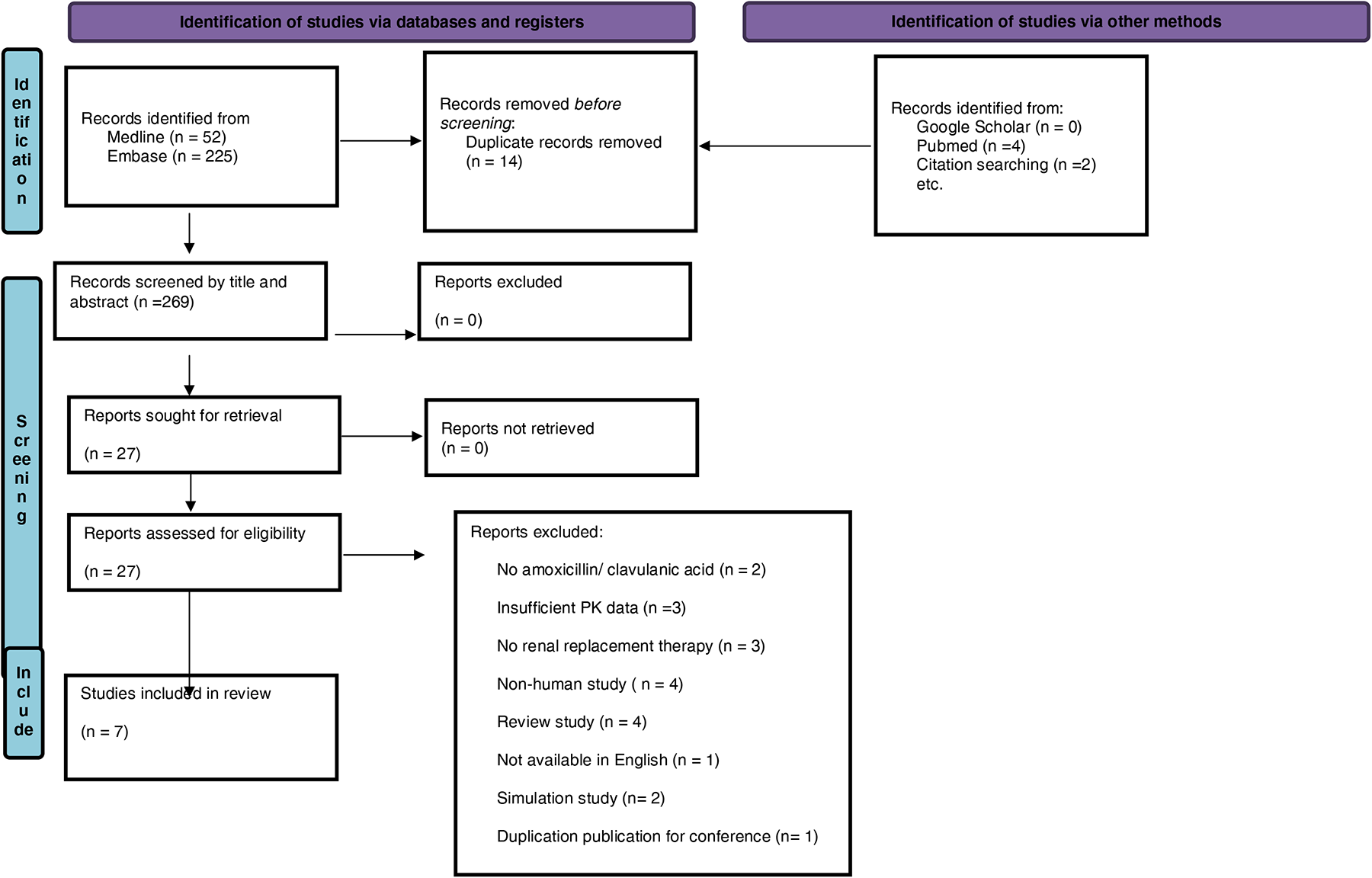
Prisma flow chart for study inclusion.

### 3.3 Data Extraction

Two authors (SAM & JP) independently extracted and collected the data in a Microsoft Excel document with pre-defined parameters. These parameters were updated after reviewing the literature to include identified relevant parameters and a repeated data extraction process for all the studies. The extracted data included the following: study design, study size, and setting, patient characteristics (age, weight, creatinine clearance, urine output), indication for antibiotic, funding source, drug name, drug dose, drug route, sampling site, sampling strategy, sample analysis, assay method, pharmacokinetic software, renal replacement mode, renal replacement machine, filter type, filter size, duration of renal replacement, blood flow rate, dialysate flow rate, pre-dilution rate, ultrafiltrate rate, delivered dose, total clearance, clearance via RRT, non-renal clearance, area under the curve, half-life, Vd, pharmacodynamic targets and any other PKPD data reported.

In studies that investigated ticarcillin and clavulanic acid [12, 13, 14], the pharmacokinetic data for ticarcillin was not extracted as this drug is not included in this systematic review, the data for clavulanic acid only was extracted. In studies that included subjects undergoing peritoneal dialysis [11] their pharmacokinetic data for Vd and clearance was not extracted as that data was aggregated. Data on the clavulanic half-life was extracted for haemodialysis subjects in this later study [11]. .

### 3.4 Quality Assessment

Two authors (SAM & JP) carried out the quality assessment independently, a third author (DL) was available to resolve disagreement. The quality assessment was done by using two scoring systems, one assessing renal replacement therapy called the acute dialysis quality initiative (ADQI) [14] and one assessing pharmacokinetics-pharmacodynamics called the quality of evidence (QoE) [15]. Based on these assessments a final aggregated quality score was allocated as low, medium, or high quality. Appendix 2 shows the details of the quality assessment scoring, Table 1 shows the final aggregate quality assessment score of the studies included.

**Table 1:** Quality assessment score of studies included.

| **Study** | **Aggregate quality assessment** |
| --- | --- |
| Dalet et al. [13] | Low quality |
| Davies et al. [19] | Low quality |
| De Schuyter et al. [18] | Medium quality |
| Hardin et al. [12] | Low quality |
| Lonsdale et al. [16] | Medium quality |
| Slaughter et al.[17] | Low quality |
| Watson et al. [11] | Low quality |

### 3.5 Data analysis

We provide a narrative systematic review of the studies. To harmonise the data, the units for drug concentration were converted from mcg/mL to mg/L, clearance was converted from mL/min to L/h for uniformity. The unit for drug dosage was converted from mg to g. The units for RRT settings were also standardised the blood flow rate was calculated as mL/min, dialysate flow as mL/h and post-dilution as mL/h. The mean if reported was represented for patient characteristics (weight, age) as well as pharmacokinetic parameters (half-life, clearance, Vd). If the mean was not reported in the individual studies it was calculated using the study data published and tabulated.

Data on amoxicillin and clavulanic acid dosages were separated into individual components. For co-amoxiclav 1.2g this was represented as amoxicillin 1g and clavulanic acid 0.2g and for ticarcillin-clavulanic acid 3.2g this was represented as ticarcillin 3g and clavulanic acid 0.2g.

We calculated the mean and standard deviation of the total clearance, RRT clearance and Vd, it was not feasible to carry out a meta-analysis on the PK data due to the clinical heterogeneity observed.

## 4 Results

### 4.1 Search and Included Studies

The search yielded 283 records, after removing duplicates there were 269 records. The titles and abstracts of the 269 records were screened, 23 full texts were reviewed for eligibility for this systematic review, of which seven articles were included. Fig. 1 shows the PRISMA flow chart for study inclusion. The publication year of the studies ranged from 1984 to 2021 with a gap in lack of publications between 1995 to 2019; Fig. 2 shows the publication year for the included studies. The characteristics of the included studies are shown in Table 2.

**Fig 2.**
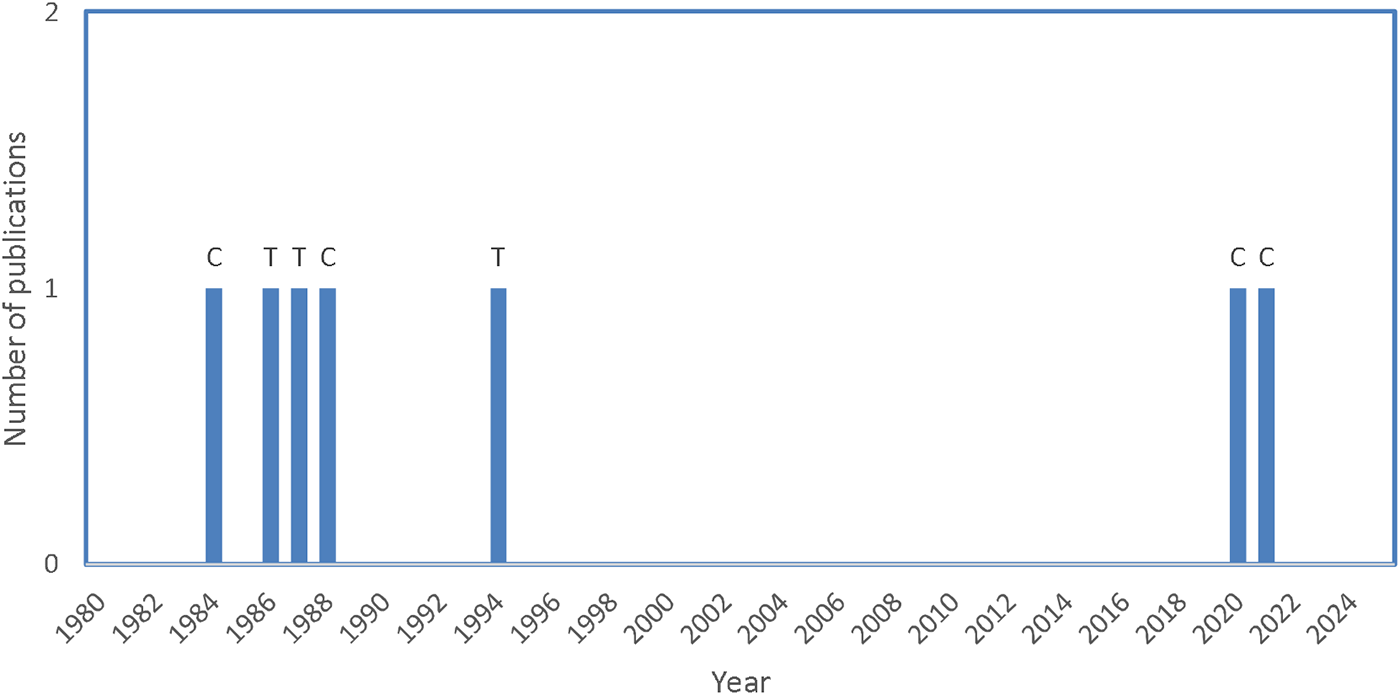
Publications on pharmacokinetics of amoxicillin and clavulanic acid in adult patients on renal replacement therapy C= co-amoxiclav T=ticarcillin-clavulanic acid

**Table 2:** Characteristics of included studies.

| Study | Country | Study design | Study size and setting | Patient characteristics |  |  |  |  | Drug name | Primary aim | Funding |
| --- | --- | --- | --- | --- | --- | --- | --- | --- | --- | --- | --- |
|  |  |  |  | Age (years) | Weight (kg) | Creatinine clearance (mL/min) | Urine output (mL/24h) | Infection |  |  |  |
| Dalet et al. [13] | Spain | Experimental pharmacokinetic study | n=5 patients on haemodialysis out of the study sample of 25 patients with renal impairment | 44 | 61 |  |  | no infection | Ticarcillin with clavulanic acid | PK of ticarcillin and clavulanic acid | not stated |
| Davies et al. [19] | United Kingdom | Experimental PK study | n=6 patients (4 male, 2 female) on regular HD | 48-65 | 46-79.5 |  |  |  | Co-amoxiclav | PK of amoxicillin and clavulanic acid during haemodialysis | Beecham Pharmaceuticals |
| De Schuyter et al. [18] | Belgium | Single-centre prospective cohort study | n=26 patients on regular HD & prescribed co-amoxiclav | 71 | 74 |  | <500, n=19<br>≥500, n=7 |  | Co-amoxiclav | population PK model for amoxicillin and clavulanic acid on intermittent high-flux HD | Fresenius Medical Care |
| Hardin et al. [12] | U.S.A | Randomised, crossover, controlled PKPD study | n=6, all male patients on chronic haemodialysis | 57 | 65.2 |  | <100 | no infection | Ticarcillin-clavulanic acid | Evaluate the pharmacodynamic/antibacterial activity of ticarcillin-clavulanic acid | National Institutes of Health-funded general clinical research unit in a Veterans Administration Medical Centre |
| Lonsdale et al. [16] | Australia | Case report | n=1 patient; adult critical care unit patient on renal replacement therapy for drug overdose, no renal impairment | 44 | 120 | 130 |  | Aspiration pneumonia; *Staphylococcus aureus* & *Klebsiella pneumoniae* in sputum; sensitive to co-amoxiclav | Co-amoxiclav | Investigating the PKPD of amoxicillin during CVVHDF | Department of intensive care, Royal Brisbane and Women's Hospital, Brisbane, Australia |
| Slaughter et al. [17] | U.S.A. | Case report | Patient n=1, chronic haemodialysis 3 times weekly in renal clinic | 16 |  |  | <100 mL | Systemic *Nocardia asteroides* infection | Co-amoxiclav | To evaluate the effects of haemodialysis on the PK of amoxicillin and clavulanic acid | Not stated |
| Watson et al. [11] | United Kingdom | Experimental pharmacokinetic study | n=3 patients out of total of 34 patients were on haemodialysis (15 female, 19 male) | 49.3 | 65.7 |  |  | no infection | Ticarcillin with clavulanic acid |  | not stated |

Abbreviations: CVVHDF (continuous venovenous haemodiafiltration); HD (Haemodialysis); PK (pharmacokinetic); PKPD (pharmacokinetics-pharmacodynamics); Vd (Volume of distribution); HD (Haemodialysis)

### 4.2 Study Population

The seven studies included had three experimental PK studies, two case reports, one single centre prospective cohort study and one randomised crossover controlled PKPD study. In one study [18] Co-amoxiclav was indicated for aspiration pneumonia where Staphylococcus aureus and Klebsiella pneumonia were grown in the sputum, they were reported as sensitive to co-amoxiclav. In one study [17] co-amoxiclav was indicated for a systemic

Nocardia asteriodes infection; one study [18] did not report the indication and the experimental studies [11, 12, 13, 19] did not have an indication for the antibiotic. Four studies investigated co-amoxiclav and three studies investigated ticarcillin-clavulanic acid. The sample size for the studies included ranged from 1 to 26 subjects and the age range was from 16 to 71 years old.

### 4.3 Renal Replacement Therapy Settings

Six of the studies investigated patients with established chronic renal impairment needing intermittent RRT over 4 h duration per session. In one study [16] the indication for RRT was for management of a drug overdose and the patient had residual renal function with an estimated creatinine clearance of 130mL/min, CVVVHDF was carried out over 192 h. One study [18] investigated 7 patients on RRT with a urine output of ≥ 500mL/24h and 19 patients with a urine output of <500mL/24h. One study [16] investigated patients on CVVHDF while one study investigated intermittent HD and PD [11], one study investigated intermittent HD and HDF [18], and three studies investigated intermittent HD [12, 13, 17, 19]. Different types of dialysers and different filters were used in each study as shown in Table 3. The blood flow rate reported ranged from 190 to 260 mL/h, the dialysate flow rate reported ranged from 8.3 to 30 mL/h. The study investigating CVVHDF [16] had an ultrafiltration flow rate of 20mL/kg/h, a delivered dose of 34 mL/kg/h and the pre-dilution was 1800 mL/h.

**Table 3:** Characteristics of renal replacement therapy in studies included.

| Study | Drug | Renal replacement therapy settings |  |  |  |  |  |  |  |  |
| --- | --- | --- | --- | --- | --- | --- | --- | --- | --- | --- |
|  |  | Renal replacement mode | Type of dialyzer | Filter type | Duration of renal replacement therapy (h) | Blood flow rate (mL/min) | Dialysate flow rate (mL/h) | Pre-dilution (mL/h) | Ultrafiltrate flow rate | Delivered dose |
| Dalet et al. [13] | Ticarcillin with clavulanic acid | Group E = haemodialysis |  | capillary cuprophane, 1.1m2 | 4 |  | 8.3 |  |  |  |
| Davies et al. [19] | Co-amoxiclav | Haemodialysis | Belco Unimat | Belco BL 612-M hollow fibre, 1.2m2 | 4 |  |  |  |  |  |
| De Schuyter et al. [18] | Co-amoxiclav | HD=12 (43%); HDF predilution=2 (7%); HDF post dilution = 14 (50%) | FX 800 Cordiax n=16; FX 600 Cordiax n=2; Evodia 1.3 n=3; Polyflux 170H n=3; Solacea 19H n=2; Theranova 400 n=2 |  | 4 | 260 |  |  |  |  |
| Hardin et al. [12] |  | Haemodialysis |  |  |  |  |  |  |  |  |
| Lonsdale et al. [16] | Co-amoxiclav | CVVHDF |  |  | 192 |  |  | 1800 | 20 (mL/kg/h) | 34 (mL/kg/h) |
| Slaughter et al. [17] | Co-amoxiclav | Haemodialysis | C-DAK | 1.8m2 | 4 | 190 | 9.5 |  |  |  |
| Watson et al. [11] |  | Haemodialysis | Gamkro AK10 | Travenol hollow-fibre (15/11) | 4 | 200 | 30 |  |  |  |

Abbreviations: HD (haemodialysis); HDF (haemodiafiltration); CVVHDF (continuous venovenous haemodiafiltration)

### 4.4 Antibiotic Treatment

Four studies investigated the combination drug co-amoxiclav (amoxicillin-clavulanic acid) and three studies investigated the combination of ticarcillin-clavulanic acid. For co-amoxiclav the routes investigated were oral and intravenous (IV) as indicated in Table 4. All the studies investigating ticarcillin-clavulanic acid used the IV route. A loading dose of 1.2g was administered in one study investigating co-amoxiclav [18], three studies investigated a single dose of ticarcillin-clavulanic; different maintenance doses were administered in four studies as described in table 4. The amoxicillin component of the dosages ranged from 500mg to 1g, the clavulanic component of the dosages investigated ranged from 100mg to 400mg as detailed in Table 4.

**Table 4:**
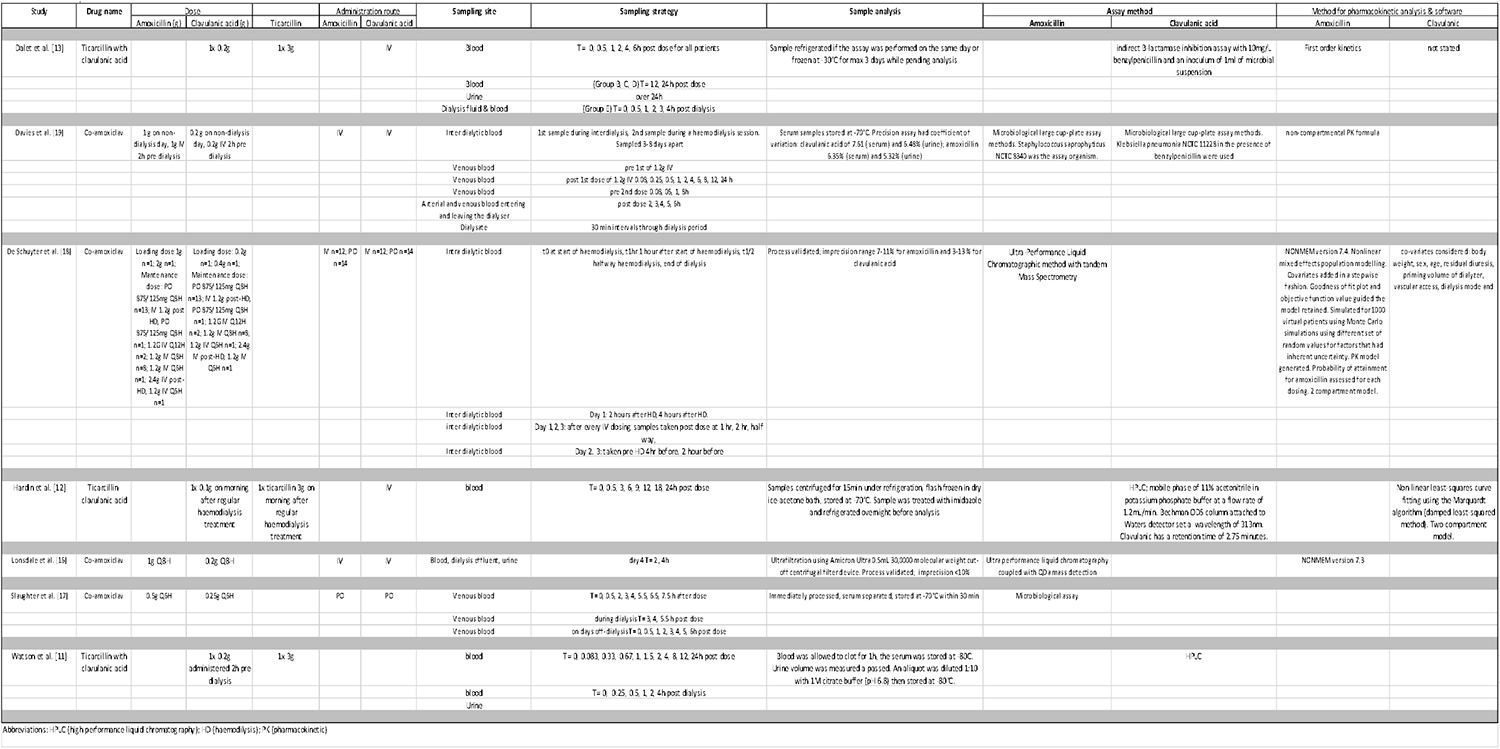
Pharmacokinetic parameters of included studies.

### 4.5 Sampling Method

All studies collected blood, five collected venous blood, one study sampled arterial and venous blood entering and leaving the dialyser, two studies sampled inter dialytic blood, one study sampled intra dialytic blood, three studies sampled dialysate fluid, one study sampled urine and effluent fluid. Most studies took the samples pre-dose, post-dose, before the RRT and after the RRT as detailed in table 3.

One study [17] analysed the exact amount of IV co-amoxiclav that was administered, the drug administration bag and IV giving set were collected after the dose was given. The remaining volume in the respective drug bag and giving set was measured to calculate the actual dose administered to the patient.

### 4.6 Analysis of sample

Samples were centrifuged and stored at temperatures ranging from -30 to -80 °C. Three studies carried out microbiological large cup plate assays, two studies carried out ultra-performance liquid chromatography and two carried out high-performance liquid chromatography. Details of validation and precision were provided in three studies [16, 18, 19] as described in Table 4.

### 4.7 Pharmacokinetic analysis method

The studies provided descriptive analysis of the pharmacokinetic data. First-order pharmacokinetic equations were used to calculate parameters for: clearance, Vd, AUC and half-life. One study applied non-linear least square curve fitting using the Marquardt algorithm [12], two studies used pharmacokinetic software NONMEM® [16, 18] for population modelling.

### 4.8 Pharmacokinetic Results

Table 5 shows the pharmacokinetic data extracted from the studies included. De Schuyter et al. [18] used a 2-compartment pharmacokinetics model, they reported the clearance of amoxicillin was 0.72 L/h for patients with <500mL/ day residual diuresis and 1.32 L/h for patients with >500mL/day residual diuresis. The Vd for amoxicillin in the central compartment was 11.3L and 12.5L in the peripheral compartment. The Vd for clavulanic was 16.2L in the central compartment and 7.4L in the peripheral compartment respectively. Other pharmacokinetic data for amoxicillin reported in this study were reported as follows: absorption rate constant was 0.78h^-1^; bioavailability was 65.1%; intercompartmental clearance between central and peripheral compartments 180mL/min; extraction ratio 74 with blood flow rate 300 mL/min; interdialytic terminal half-life was reported as 10.8h for patients with <500mL residual diuresis and 6.1h for patients with >500mL residual diuresis; the intradialytic half-life for patients with <500mL residual diuresis was 0.56h and 0.53h for patients with >500mL residual diuresis. The other pharmacokinetic data reported for clavulanic in this study were as follows: absorption rate constant was 1.14h^-1^; bioavailability was 37%; intercompartmental clearance between central and peripheral compartments 66.7mL/min; extraction ratio 78 at a blood flow rate of 300 mL/min; interdialytic terminal half-life was reported as 2.9h; the intradialytic half-life was 0.62h.

**Table 5:**
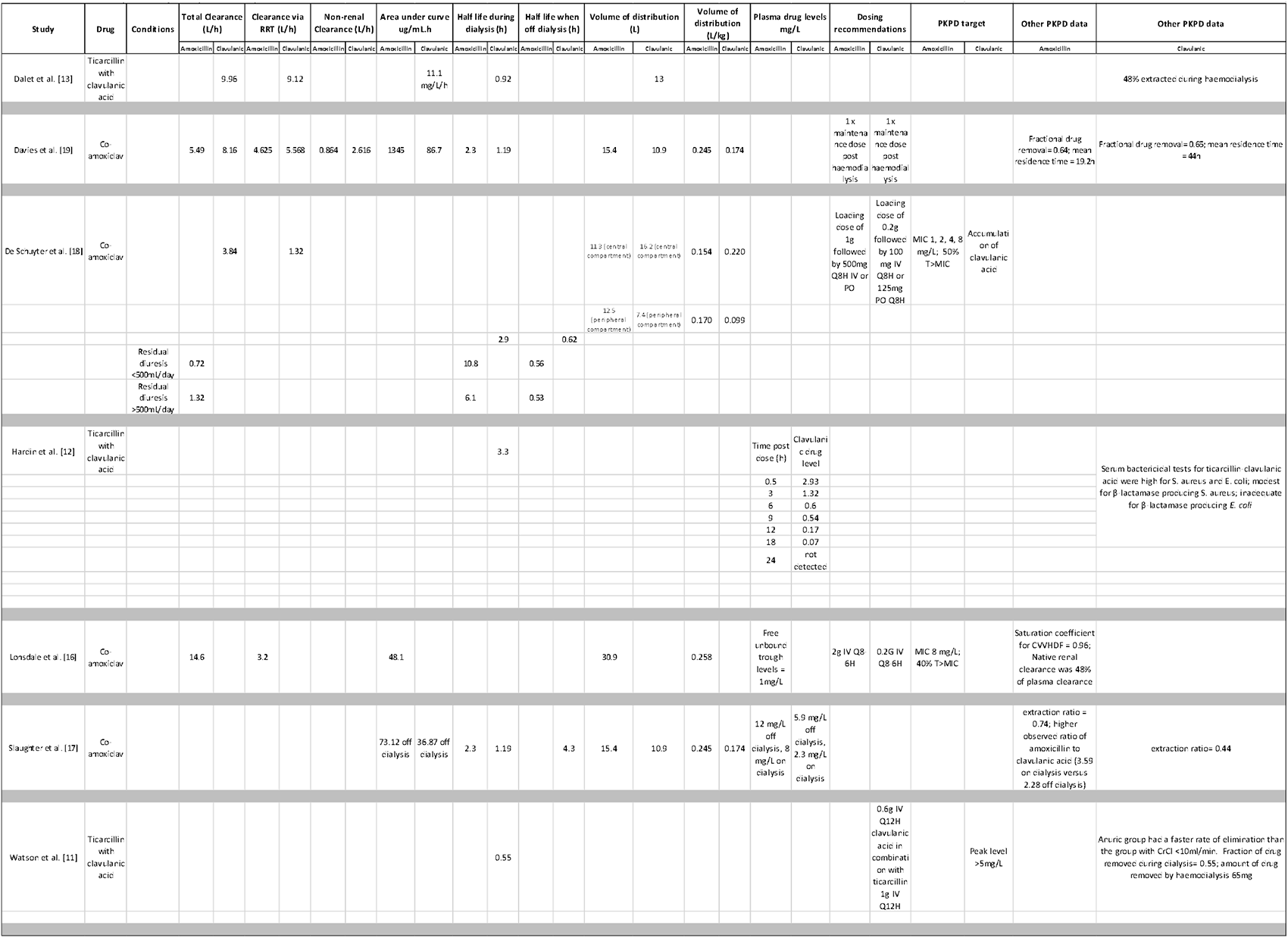
Pharmacokinetic and pharmacodynamic parameters reported in studies.

Lonsdale et al. [16] reported the total clearance of amoxicillin as 14.6 L/h and the clearance via renal replacement therapy was 3.2 L/h. The Vd was reported for amoxicillin as 30.9L for 120kg patient. The native kidney was suggested to contribute to 48% of the plasma drug clearance for amoxicillin.

Davies et al. [19] reported the total clearance of amoxicillin to be 5.49 L/h and clavulanic clearance to be 8.16 L/h. They also reported the clearance via renal replacement therapy to be 4.63 L/h for amoxicillin and 5.67 L/h for clavulanic. The Vd for amoxicillin was 15.4 L and for clavulanic 10.9L for a mean weight of 62.75kg. Elimination as a result of haemodialysis was high with 77 and 92.8 mL/min for amoxicillin and clavulanic acid respectively. A greater percentage of the amoxicillin dose was found in the dialysate compared with clavulanic acid, with means of 47.2 and 34.1 % of the dose, respectively. The terminal half-life during dialysis for amoxicillin was 2.3 h and for clavulanic acid 1.19 h.

Slaughter et al. [17] reported that there is increased clearance of both amoxicillin and clavulanic acid, the results of this study [17] show the lower serum concentrations of both drugs during dialysis compared to the off-dialysis period. For amoxicillin, the drug concentrations were 8.0 +/-1.8 vs 12.4 +/-0.9 mg/ml and for clavulanic acid, the drug concentrations were 2.3 +/- 0.8 vs 5.9 +/-1.2 mg/ml, respectively. Clavulanic acid was reported to have a high extraction ratio of 0.74 compared to amoxicillin at 0.44. The half-life of clavulanic acid during the off-dialysis period was 4.3 h and the area under the curve was 36.87 mg/mL.h. The half-life of amoxicillin was not attained, however, the area under the curve was 73.13 mg/mL.h on the off-dialysis day.

Dalet et al. [13] reported for clavulanic acid the following parameters: clearance 9.12 L/h via haemodialysis; total clearance 9.96 L/h; area under curve 11.1 mg/mL.h; half-life 0.92; Vd 13L; 48% of clavulanic acid was extracted during haemodialysis.

Watson et al [11] reported for clavulanic acid a half-life of 0.55h and Hardin et al. [12] reported a half-life of 3.3h.

Pharmacodynamic target and co-amoxiclav dosing recommendations:

Davies et al. [19] recommended a supplemental maintenance dose of co-amoxiclav post haemodialysis. The dose was not specified and the target pharmacodynamic parameter for co-amoxiclav was not stated. De Schuyter et al. [18] had an amoxicillin target MIC of 1, 2, 4, 8mg/L for 50% fT>MIC; they recommend a loading dose of co-amoxiclav 1.2g IV followed by a maintenance dose of 600mg IV Q8H or 625mg PO Q8H. Lonsdale et al. [16] recommended a dose of co-amoxiclav 2.2 g IV Q6 or 8H for a target of 40% fT>MIC of 8mg/L. Watson et al. [11] aimed for a peak clavulanic acid level of >5mg/L, they recommended a dose of clavulanic acid 600mg IV Q12h.

### 4.9 Strength of Evidence

The quality assessment of the included studies is shown in Table 1 and Appendix 2. The strength of evidence was of medium quality for two studies [16, 18] and low quality for five studies [11, 12, 13, 17, 19]. Details on reporting for RRT settings were missing in some studies with respect to the dialysis flow rate, blood flow rate, time spent on RRT, the indication for filtration, the severity of illness and patient outcomes. Two studies were case studies with PK, four studies were individual PK studies with no simulation and two studies were PK studies with simulation and modelling using PKPD software.

## 5 Discussion

The included pharmacokinetic studies on co-amoxiclav spanned between the years 1984 to 2021, with a publication gap between 1995 to 2019. Seven studies identified during this period illustrate the scarcity of studies. The quality assessment score for the included studies showed the studies were of low to intermediate quality according to the aggregate quality score in Appendix 2.

There were many differences with respect to the baseline characteristics of the studies, patient demographics, the type of renal replacement therapy settings used and the drug assay methodology. Due to the heterogeneity of the data found as well as the lack of data, the pharmacokinetic data could not be pooled for further pharmacokinetic modelling to recommend optimal dosing for co-amoxiclav in critically ill patients on renal replacement therapy.

There were many confounding factors seen in the studies which could affect the pharmacokinetics of co-amoxiclav resulting in bias. The assay methods were different. Two studies [16, 18] both used ultra-performance liquid chromatography coupled with mass spectrometric detection, two studies used HPLC [12, 11], one study [17] did not clarify the assay method used while three studies [13,17,19] used microbiological assay methods. The differences in assay methods will affect the respective validity, and reliability of the pharmacokinetic data obtained in each respective study. Microbiological assays have now been superseded by HPLC and mass spectrometry. In In one study [13] the samples were refrigerated if they were assayed on the same day. The potential variability in the storage of the samples in the freezer or fridge may impact the stability of the clavulanic acid resulting in an inaccurate assay result. Data from a study assessing the stability of amoxicillin and clavulanic acid showed that amoxicillin retained 90% of its initial concentration for 80.3 h at 4°C, 24.8h at 25°C and 9h at 37°C; clavulanic acid retained 90% of its initial concentration for 152 h at 4°C, 26h at 25°C and 6.4h at 37°C [20]. Studies have recommended storing clavulanic acid at -80°C within 1 h after withdrawal due to the poor stability of clavulanic acid [21]. None of the studies reported the time period between sampling and refrigeration, or the storage period. Only one study [11] stored the samples at -80°C. The storage conditions would impact the validity of the assays performed on clavulanic acid.

The pharmacokinetic analysis differed in the studies, two studies used non-linear mixed effects population modelling software (NONMEM) [16, 18] and 5 studies provided a descriptive analysis of pharmacokinetic results, calculating non-compartmental parameters for Vd, clearance, AUC and half-life. Two studies administered oral co-amoxiclav [17, 18]. Oral co-amoxiclav bioavailability can be affected by confounding patient factors. These differences in the studies may affect the pharmacokinetic parameters calculated.

The sampling strategies used in each study varied greatly between with respect to different sampling times and different sampling sites. The renal replacement mode therapy applied to each patient was also different with haemodialysis, high flux haemodialysis and CVVHDF. Six of the studies [11, 12, 13, 17, 18, 19] carried out intermittent haemodialysis and one study [16] carried out continuous renal replacement therapy. Renal replacement therapy was carried out using different machines, filters, filter sizes, RRT fluids, dialysis flow rates and blood flow rates. These differences could potentially lead to a difference in the clearance of the drug secondary to the renal replacement method used. In addition, the patient’s intrinsic renal function will contribute towards the total clearance of the drug. This has been demonstrated in the case study [16] which demonstrated augmented renal clearance by the patient that contributed towards 48% of the amoxicillin clearance. Clavulanic acid undergoes extensive hepatic metabolism, this has not been accounted for in any of the studies included. The multiple confounding factors seen in the studies as well as the small sample size illustrate the need for further pharmacokinetic studies to ascertain how renal replacement therapy affects co-amoxiclav pharmacokinetics.

The studies investigating ticarcillin-clavulanic acid mainly yielded useful pharmacokinetic data on clavulanic half-life.

The mean pharmacokinetic results of this systematic review as shown in Table 6. The data should be interpreted with caution due to the heterogeneity observed. For amoxicillin, the mean Vd was 0.214 L/kg, standard deviation of 0.098. This Vd is much lower than the healthy volunteer (0.319 L/kg) and is also lower than that of a critically ill patient 0.317 to 0.365 L/kg. For clavulanic acid the mean Vd was 0.133L/kg, standard deviation of 0.086. This Vd is again much lower than that reported in a healthy volunteer (0.33L/kg) and a critically ill patient (0.256 to 0.301 L/kg).

**Table 6:** The mean clearance and volume of distribution of amoxicillin and clavulanic acid on included studies.

|  | Total Clearance (L/h) |  | RRT clearance (L/h) |  | Volume of distribution (L/kg) |  |
| --- | --- | --- | --- | --- | --- | --- |
|  | Amoxicillin | Clavulanic | Amoxicillin | Clavulanic | Amoxicillin | Clavulanic |
| Mean | 5.53 | 7.32 | 2.61 | 5.34 | 0.214 | 0.133 |
| Standard deviation | 6.41 | 3.15 | 1.01 | 3.91 | 0.098 | 0.086 |
| Healthy volunteer | 13.5 to 25 | 16.1 |  |  | 0.319 | 0.33 |
| Critically-ill person | 10.3 | 9.4 |  |  | 0.317 to 0.365 | 0.256 to 0.301 |

RRT (renal replacement therapy)

For amoxicillin, the mean total clearance was 5.53L/h, standard deviation of 7.32. This amoxicillin clearance is much less than that of a healthy volunteer (13.5 to 25 L/h) and that of a critically ill patient (10.3L/h). For clavulanic acid, the total mean clearance was 7.32 L/h, standard deviation of 3.15. This clavulanic acid clearance is much less than that of a healthy volunteer (16.1 L/h) and a critically ill patient (9.5 L/h).

The standard deviation shows that there is a wide variance in the pharmacokinetic data, this is highly likely due to the stark differences between studies.

To aim for a free antibiotic concentration of amoxicillin above the minimum inhibitory concentration of the target pathogen (fT>MIC) of 8mg/L Lonsdale et al. [18] recommend a dose of co-amoxiclav IV 2.2 g every 6-8 hours in addition to therapeutic drug monitoring. The dose administered in the study was 1.2g IV every 8 hours, trough concentrations of amoxicillin were reported as 1mg/L. As The EUCAST range is between 0.25 to 8 mg/L for amoxicillin, this shows that the co-amoxiclav dose was likely subtherapeutic. Lonsdale et al. [18] suggested that the infection was treated despite the subtherapeutic levels due to the possibility of the MIC being lower than 8mg/L for the respective pathogen. The MIC was not assessed in this case.

De Schuyter et al. [18] recommended a loading dose of 1.2g followed by 500mg/100mg IV Q8H or 500mg/125mg PO Q8H. Davies et al. [13] recommend giving one maintenance dose of co-amoxiclav post-haemodialysis.

The renal drug database [3] recommends a dose of 1.2g IV stat followed by 600mg Q8H or 1.2g Q12h for co-amoxiclav with haemodialysis, heamodiafiltration, and high flux haemodialysis. For continuous venovenous haemodialysis the recommended dose is 1.2g IV Q12H; this is the same dosage recommendation from the study by De Schuyter et al. [18]. Medicines complete critical illness [9] recommends for patients on continuous renal replacement including CVVHF, CVVHDF, and CVVHD a co-amoxiclav dose of 1.2g Q8H for the first 24-48 hours. If the effluent rate on day 2 is less than 25mL/kg/h a dose reduction of 1.2g Q12H is recommended. If the effluent rate is greater than 30mL/kg/h a higher dose of 1.2g Q6H is recommended. This dosage recommendation is based on the study by Lonsdale et al. [16]

The paucity of data available along with the non-uniformity of data reported limits the data analysis that can be performed. In addition, the heterogeneity of the study designs introduces limitations on what can be concluded from the data itself. It also limited the ability to perform a meta-analysis.

Limitations of this systematic review are that the Acute Dialysis Quality Initiative (ADQI) quality assessment method used to assess the studies identified has the use of co-interventions published by the authors such as antibiotics as a positive point in the grading method. By default, the inclusion criteria for this systematic review requires that all the studies selected had patients on antibiotics and renal replacement therapy, hence this marker of quality was a base line for all the studies included and did not contribute towards drawing out the strength between these respective studies. Another limitation is that studies not published in the English language were excluded.

This systematic study aimed to find what the pharmacokinetics of co-amoxiclav are in patients undergoing RRT. The studies provide limited PKPD data. The results highlight the need for further studies to assess the PKPD of co-amoxiclav in patients on renal replacement therapy.

## 6 Conclusion

Further pharmacokinetic-pharmacodynamic studies are needed to clarify primary pharmacokinetic parameters of co-amoxiclav during renal replacement therapy. More rigorously designed studies reporting co-variate factors such as assay temperature, residual kidney function and RRT characteristics; would generate more reliable and perhaps more consistent data on co-amoxiclav clearance and volume of distribution. This is required to reduced variability in clinical approaches and standardise future RRT dosing to reduce reported variability in PK target obtainment and outcomes.

## Data Availability

All data produced in the present study are available upon reasonable request to the authors

## Appendix 1: Search strategy

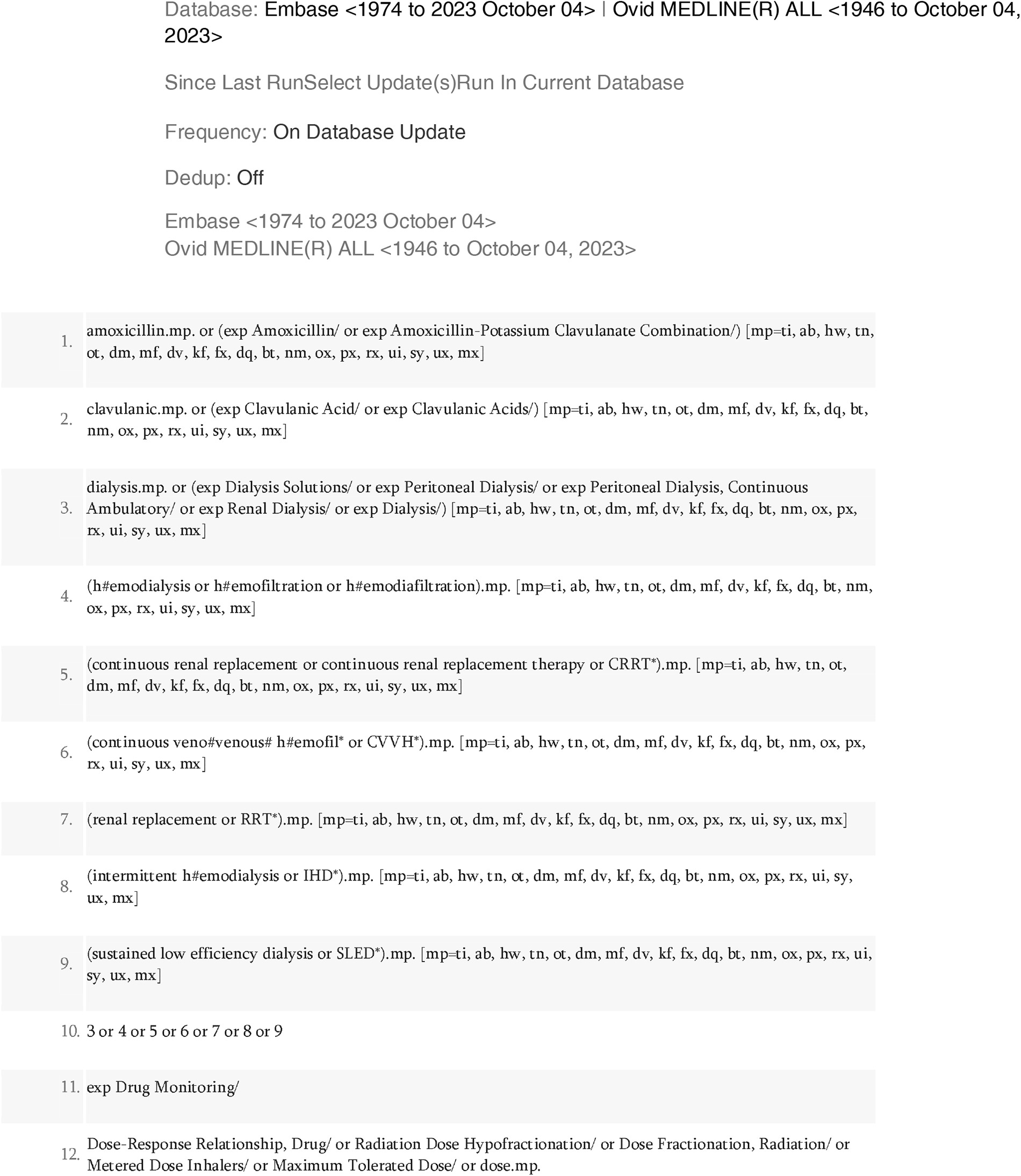
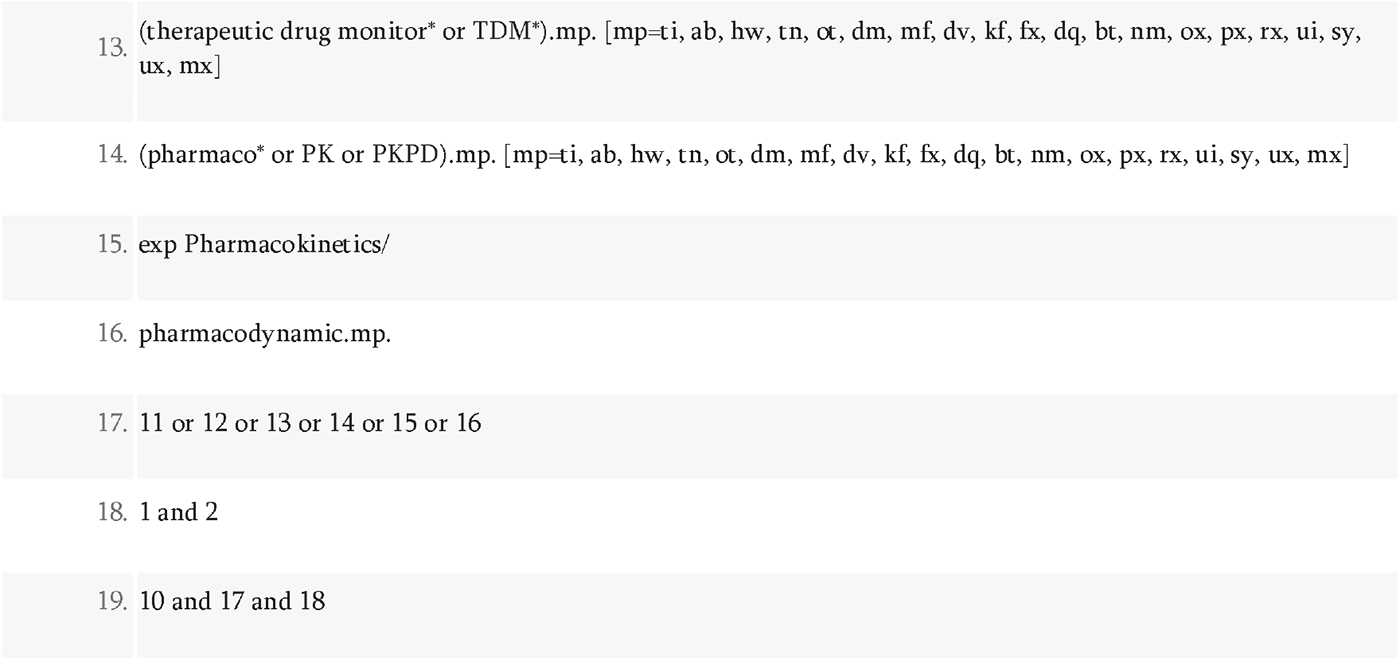

## Appendix 2: Outcome of quality assessment of studies included

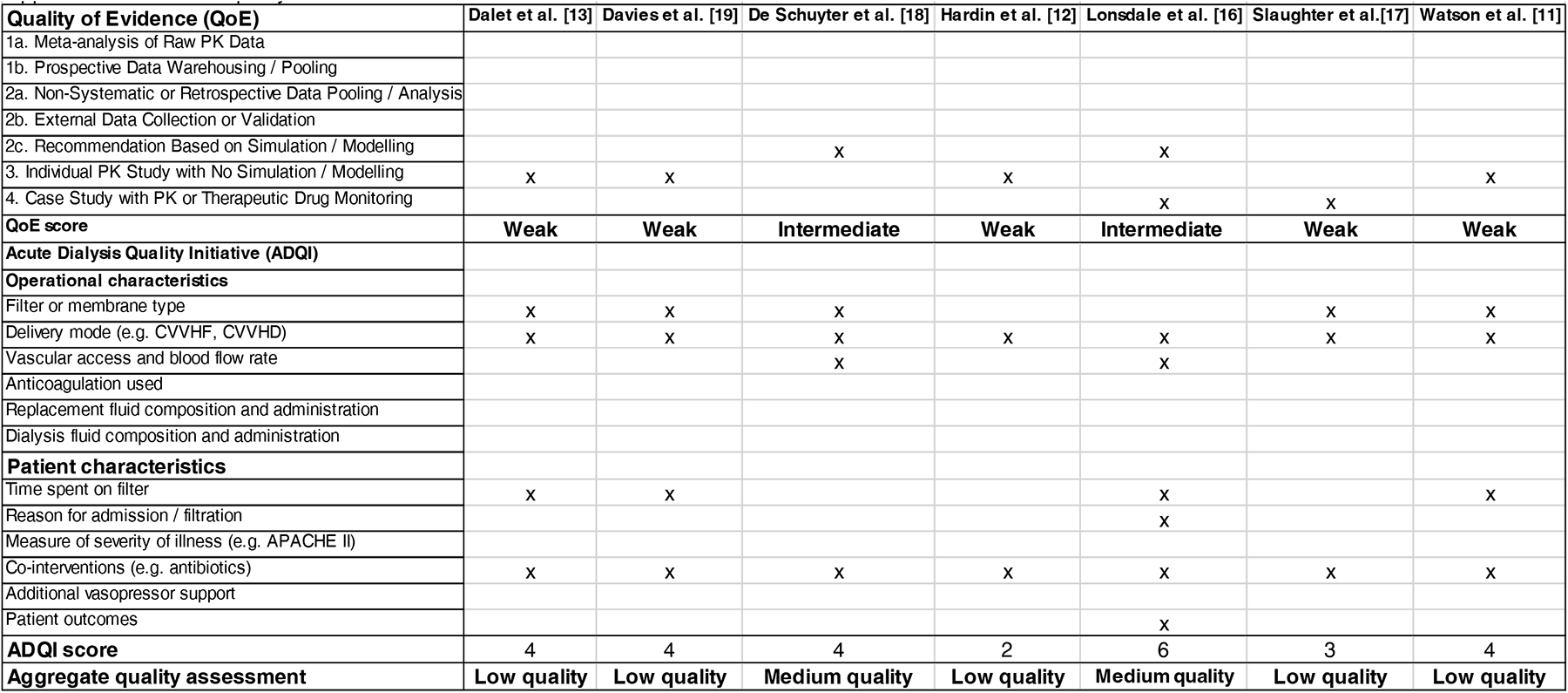
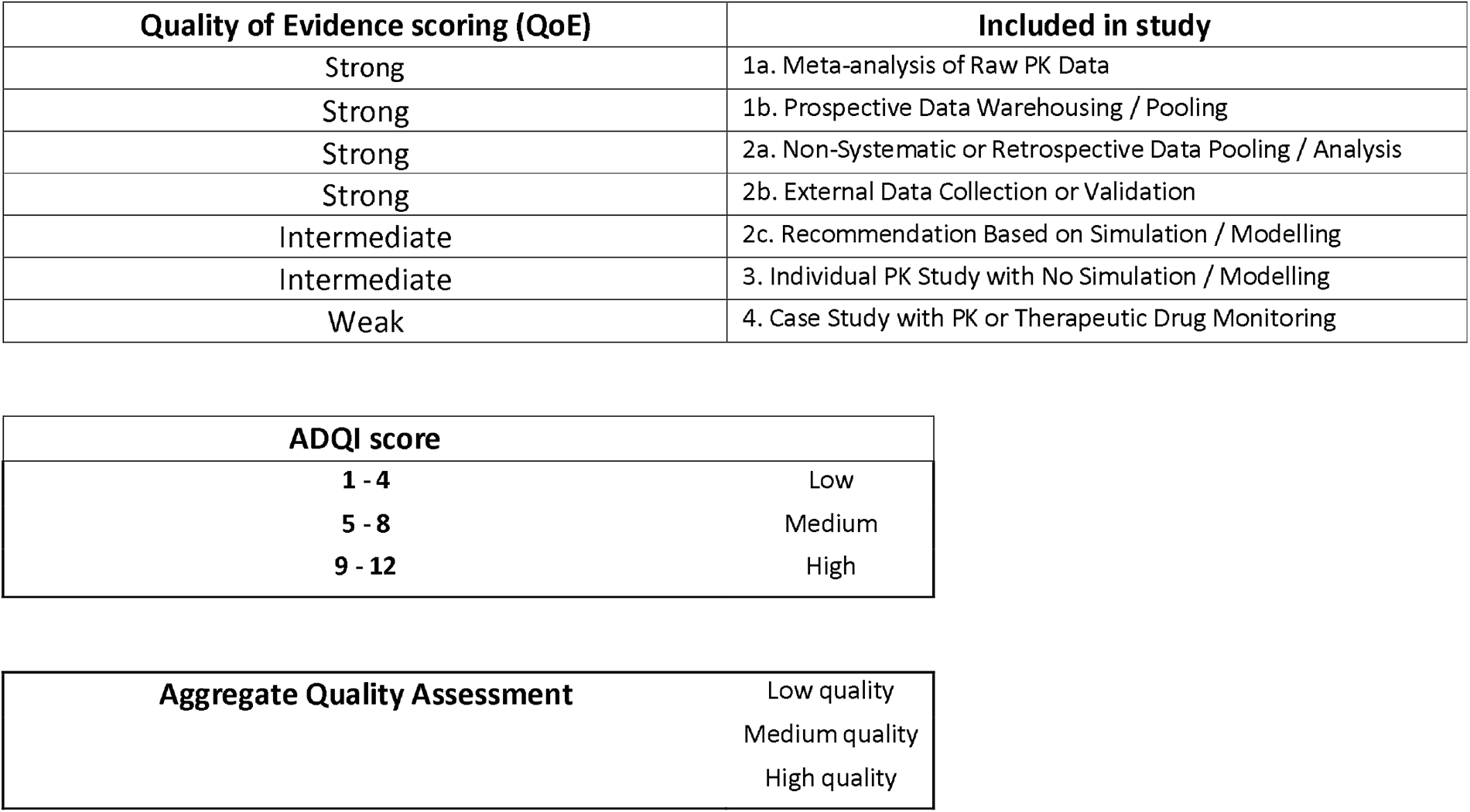

